# SARS-CoV-2 Infection-Associated Hemophagocytic Lymphohistiocytosis: An autopsy series with clinical and laboratory correlation

**DOI:** 10.1101/2020.05.07.20094888

**Authors:** Andrey Prilutskiy, Michael Kritselis, Artem Shevtsov, Ilyas Yambayev, Charitha Vadlamudi, Qing Zhao, Yachana Kataria, Shayna R. Sarosiek, Adam Lerner, J. Mark Sloan, Karen Quillen, Eric J. Burks

## Abstract

**Background:** A subset of COVID-19 patients exhibit clinical features of cytokine storm. However, clinicopathologic features diagnostic of hemophagocytic lymphohistiocytosis (HLH) have not been reported. Pathologic studies to date have largely focused on the pulmonary finding of diffuse alveolar damage (DAD). To this aim, we study the reticuloendothelial organs of four consecutive patients dying of COVID-19 and correlate with clinical and laboratory parameters to detect HLH.

**Methods:** Autopsies restricted to chest and abdomen were performed on four patients who succumbed to COVID-19. Spleen, liver, and multiple pulmonary hilar/mediastinal lymph nodes were sampled in all cases. Bone marrow was obtained by rib squeeze in a subset of cases. Routine H&E staining as well as immunohistochemical staining for CD163 was performed to detect hemophagocytosis. Clinical and laboratory results from pre-mortem blood samples were used to calculate H-scores.

**Findings:** All four cases demonstrated DAD within the lungs. Three of the four cases had histologic evidence of hemophagocytosis within pulmonary hilar/mediastinal lymph nodes. One case showed hemophagocytosis in the spleen but none showed hemophagocytosis in liver or bone marrow. Lymphophagocytosis was the predominant form of hemophagocytosis observed. One patient showed diagnostic features of HLH with an H-score of 217 while a second patient was likely HLH with a partial H-score of 145 due to missing triglyceride level. Both patients exhibited high fever and early onset rise in serum ferritin; however, neither bicytopenia, pancytopenia, nor hypofibrinogenemia were observed in either. The remaining two patients had H-scores of 131 and 96.

**Interpretation:** This is the first report of SARS-CoV-2 associated HLH. Identification of HLH in a subset of patients with severe COVID-19 will inform clinical trials of therapeutic strategies.

## Introduction

The global pandemic of severe acute respiratory syndrome coronavirus 2 (SARS-CoV-2) has infected over 3.5 million people worldwide and led to more than 250,000 deaths within just 5 months of the initial outbreak in Wuhan, China.^1^ The rapid spread of the virus is largely the result of community transmission by individuals who are asymptomatic.^2^ While some individuals remain asymptomatic throughout the disease course^3^ others develop a flu-like illness with viral pneumonia which may progress to acute respiratory distress syndrome, viral sepsis^4^, and cytokine storm.^5^ The latter has raised clinical concerns for hemophagocytic lymphohistiocytosis (HLH) and the overlapping disorder, macrophage activation syndrome (MAS), highlighting the potential for therapeutic cytokine blockade.^6–8^

Despite these clinical observations, there has been relatively little clinicopathologic correlation with the post-mortem findings in COVID-19 which to date have generally focused on the pulmonary histology of diffuse alveolar damage.^9–12^ Herein we report the first cases of SARS-CoV-2 associated HLH diagnosed post-mortem using histologic, clinical and laboratory criteria.

## Materials and Methods

After obtaining consent from next of kin, autopsy was performed on four patients testing positive for SARS-CoV-2 nucleic acid on antemortem upper respiratory swab. Chart review of the electronic health record was performed to determine age, gender, race, date of symptom onset, date of hospitalization, date and types of treatment, CBC parameters, serum fibrinogen, ferritin, aspartate aminotransferase (AST), and C-reactive protein (CRP). Laboratory values were obtained for each case on the day of ICU transfer (average of hospital Day 5 for patient 1, 3, 4) and on hospital day 5 for patient 2. Ferritin levels were recorded over entire hospital course. Triglyceride studies were performed on available antemortem blood samples after death in 3 of 4 patients.

The autopsies were performed in a negative pressure room using Personal Protective Equipment (PPE) including N95 mask and powered air purifying respirator (PAPR). Autopsies were limited to chest and abdomen. Harvested organs were thinly sliced and fixed for 24 hours in 10% neutral buffered formalin (NBF). Tissue blocks were prepared and fixed for an additional 6-12 hours in 10% NBF before processing. Bone marrow obtained by rib squeeze was fixed for 12-36 hours in B-Plus Fix™ (BBC Biochemical) fixative prior to processing without decalcification. Hematoxylin and Eosin (H&E) stained sections were prepared and reviewed by two board certified anatomic pathologists (EB and QZ) one of whom is a board-certified hematopathologist (EB) with familiarity in evaluating samples for hemophagocytosis.

Immunohistochemistry and in situ hybridization were performed using freshly cut 5 μm thick FFPE tissue sections from lymph nodes, spleen, liver, and bone marrow. Slides were stained with CD163 clone MRQ-26 (Ventana, 760-4437), HHV-8 clone 13B10 (Leica, NCL-L-HHV8-LNA), CMV clone DDG9/CCH2 (Cell Marque, 213M-18), and EBER 1 DNP Probe (Ventana, 760-1209A) on a Ventana Benchmark Ultra (Ventana Medical Systems, Tucson, AZ, USA) after onboard heat induced epitope retrieval with CC1 buffer (Ventana, 950-124) for immunohistochemistry or protease digestion for in situ hybridization. Slides were visualized with Optiview detection (Ventana, 760-700). Internal and external controls were examined for each slide and judged satisfactory before interpretation. As controls, spleen and mediastinal lymph nodes from archived autopsies of five patients with non-COVID-19 related ARDS performed between 2007-2019 were stained with CD163. The presence of histiocytes phagocytosing nucleated cells was counted in ten high power fields (200x magnification) focusing on sinusoids and the frequency of hemophagocytes containing single or multiple nucleated cells was calculated per 2mm^2^.

H-scores were calculated based upon clinical and laboratory data and using the presence of hemphagocytosis in one or more reticuloendothelial organs at autopsy to define the pathologic component of the score. The H-score is a validated tool which predicts HLH with 90% accuracy using a cutoff of 169.^13,14^

The Institutional Review Board reviewed this study and waived jurisdiction.

## Results

### Patient Characteristics and Treatment

Clinical features of the four patients are summarized in Table 1: they included three men and one woman ranging from 64-91 years of age. Three were African American while one was Caucasian. Disease course included progressive hypoxia leading to intubation in two patients who died on day 15 and day 18 after symptom onset while the remaining two patients were not intubated according to do not intubate orders. All patients received antibiotics during the hospital course including azithromycin, doxycycline, and/or ceftriaxone. Hydroxychloroquine was administered to two patients. Sarilumab, a human monoclonal antibody against the interleukin 6 receptor (IL-6R) was given to two patients and Anakinra, an interleukin 1 receptor (IL-1R) antagonist was given to one patient.

**Table 1:** Clinicopathologic Features.

|  | Patient 1 | Patient 2 | Patient 3 | Patient 4 |
| --- | --- | --- | --- | --- |
| Age | 72 | 91 | 72 | 64 |
| Gender | Male | Male | Male | Female |
| Race | CA | AA | AA | AA |
| Symptom Onset Until |  |  |  |  |
| Hospitalization (days) | 4 | 1 | 3 | 5 |
| Treatment (days) | D5-HCQ/AZI<br>D6,7,8-Anakinra<br>D7-Intubation | D1-HCQ/DOX/AZI | D3-CRO/AZI<br>D4-Sarilumab | D5-Sarilumab<br>D6-CRO<br>D12-Intubation |
| Death (days) | 18 | 8 | 6 | 15 |
| Highest body temperature (°F) | 104 | 102.8 | 101.3 | 102.7 |
| White blood cells count (K/uL) | 6.2 | 10.6 | 11.1 | 9.8 |
| Absolute lymphocyte count (K/uL) | 0.6 | 0.8 | 0.8 | 1.2 |
| Hemoglobin (g/dL) | 10.2 | 8.9 | 10.5 | 14.6 |
| Platelet count (K/uL) | 108 | 317 | 681 | 261 |
| Fibrinogen (mg/dL) | 418 | 800 | 800 | 559 |
| Ferritin (ng/mL) | 7679 | 4095 | 167 | 397 |
| Triglyceride (mg/dL) | 1316 | .. | 162 | 353 |
| Aspartate transaminase (U/L) | 63 | 37 | 59 | 32 |
| C-reactive protein (mg/L) | 81.6 | 354.6 | 365.1 | 77.7 |
| Hepatomegaly | No | No | No | No |
| Splenomegaly | No | Yes | No | No |
| Hemophagocytosis |  |  |  |  |
| Lymph node | Yes | Yes | Yes | No |
| Spleen | No | Yes | No | No |
| Liver | No | No | No | No |
| Bone Marrow | .. | .. | No | No |
| **H-score** | **217** | **145\*** | **131** | **96** |
| **HLH Syndrome** | **Definite** | **Probable** | **Absent** | **Absent** |

\*Partial H-score as triglyceride level was not available for this patient. Laboratory values were obtained for each case on the day of ICU transfer (average of hospital Day 5 for patient 1, 3, 4) and on hospital day 5 for patient 2.
Abbreviations: CA, Caucasian; AA, African-American; HCQ, hydroxychloroquine; AZI, azithromycin; DOX, doxycycline; CRO, ceftriaxone; .., not done.

### Autopsy Findings

The lungs showed acute exudative phase of DAD in all four patients consistent with the clinical presentation of acute respiratory distress syndrome (ARDS).

Mediastinal and pulmonary hilar lymph nodes were grossly enlarged and contained clusters of hemophagocytic histiocytes in 3 of 4 cases. In patient 1 this was characterized by a marked distention of cortical and subcortical sinuses with focal necrosis (Figure 1A/B) as well as lymphocyte depletion. In patients 2 and 3 the finding was more limited with multifocal clusters of hemophagocytic histiocytes localizing in the subcapsular sinuses (Figure 1C/D). Lymphoid depletion was not identified in these cases; follicular and interfollicular hyperplasia was present in patient 3 (images not shown). Lymphophagocytosis was the predominant form of hemophagocytosis in all patients (Figure 1E) with frequent occurrence of multiple lymphocytes engulfed within single histiocytes most easily visualized using CD163 immunohistochemical stain (Figure 1F). Immunohistochemistry for human herpes virus-8 (HHV-8), cytomegalovirus (CMV), and Epstein-Barr virus by in situ hybridization for Epstein-Barr virus small RNA (EBER) were negative in lymph nodes with hemophagocytosis (images not shown). As controls, mediastinal lymph nodes from autopsies of five patients with non-COVID-19 related ARDS were stained with CD163 and only rare lymphophagocytic histiocytes were observed (range, 0-0.6 per 2mm^2^) and no histocytes containing multiple lymphocytes were seen (images not shown).

**Figure 1.**
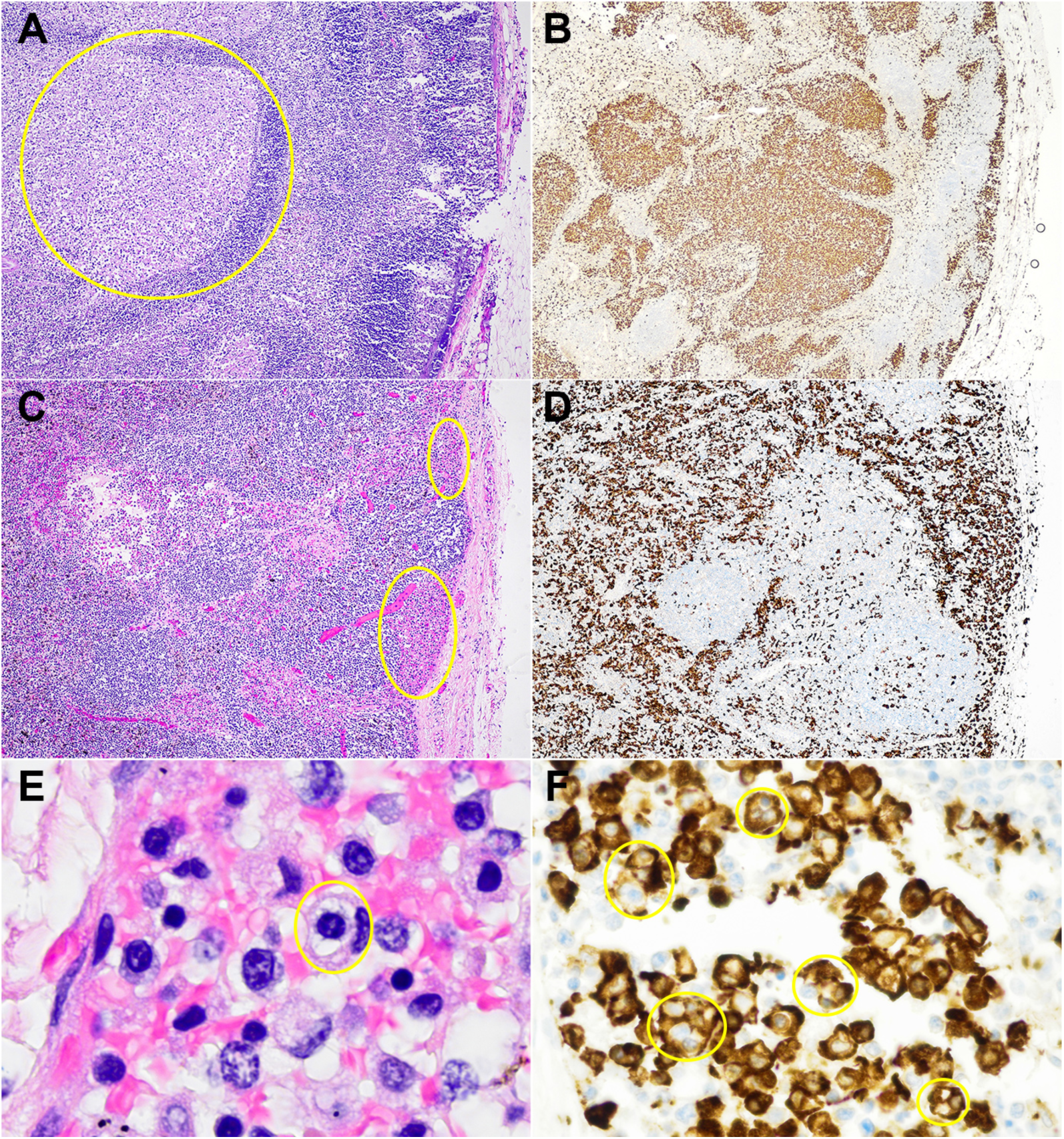
Hemophagocytosis in pulmonary hilar and mediastinal lymph nodes. (A/B) H&E and CD163 immunohistochemical stains of lymph node from patient 1 showing distended cortical and subcortical sinusoids filled with histiocytes exhibiting focal necrosis. (C/D) H&E and CD163 immunohistochemical stains of a lymph node from patient 2 showing a lesser degree of sinusoidal expansion, predominantly filling the subcapsular sinuses. Hemophagocytosis consisted predominantly of lymphophagocytosis in all cases and seen on (E) H&E stain and highlighted with (F) CD163 immunohistochemical stain where numerous histiocytes phagocytosing one to several lymphocytes were apparent.

The spleen was enlarged in a single patient (patient 2) with a soft and friable gross appearance which ruptured upon evisceration. Microscopically there were numerous areas of red pulp hemorrhage with admixed phagocytic histiocytes, focally showing hemophagocytosis (Figure 2A-C) and white pulp depletion. Spleen size was normal in the remaining patients. In patient 1 there was white pulp depletion with red pulp infarction (Figure 2D), histiocytic hyperplasia (Figure 2E), and numerous hemosiderin-laden macrophages suggestive of prior red cell phagocytosis (Figure 2F). In patient 3 and 4, the spleens showed normal to slightly hyperplastic white pulp with red pulp congestion but lacking hemophagocytosis. As controls, sections of spleen from autopsies of five patients with non-COVID-19 related ARDS were stained with CD163 and only rare lymphophagocytic histiocytes were observed (range, 0-0.6 per 2mm^2^) and no histocytes containing multiple lymphocytes were seen (images not shown).

**Figure 2.**
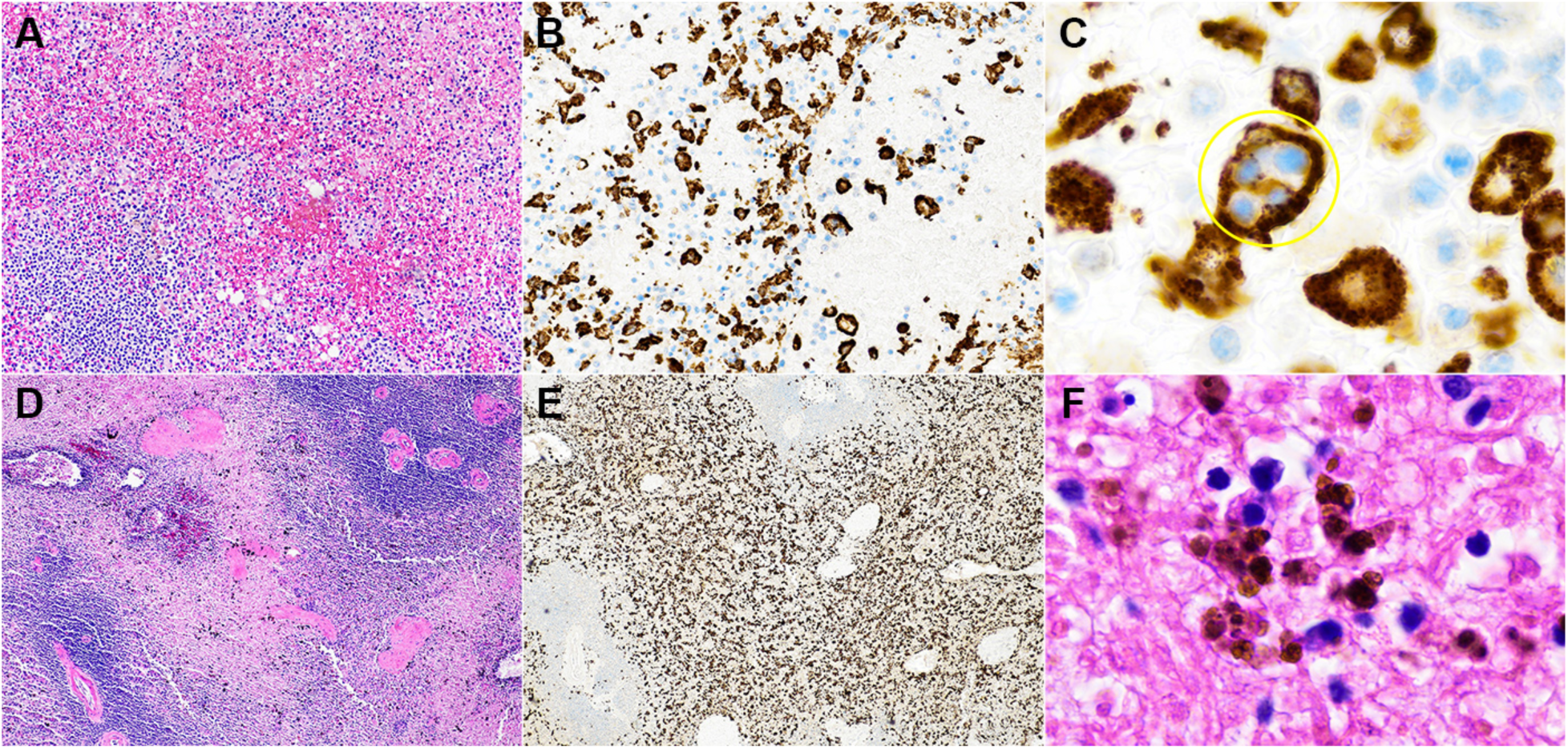
Hemophagocytosis in spleen. (A) H&E stain of spleen from patient 2 with white pulp depletion and red pulp hemorrhage with (B) CD163 immunohistochemical stain demonstrating mild histiocytic hyperplasia and (C) lymphophagocytosis. (D) H&E stain of spleen from patient 1 showing mild white pulp depletion and red pulp infarction with (E) CD163 immunohistochemical stain showing moderate histiocytic hyperplasia and (F) hemosiderin-laden macrophages suggestive of prior erythrophagocytosis.

The liver was not enlarged in any of the patients. Histologically there was mild centrilobular congestion with mild steatosis in a subset of cases but without significant portal or lobular inflammation. Minimal Kupffer cell hyperplasia was seen on CD163 staining but hemophagocytosis was not observed (Figure 3).

**Figure 3.**
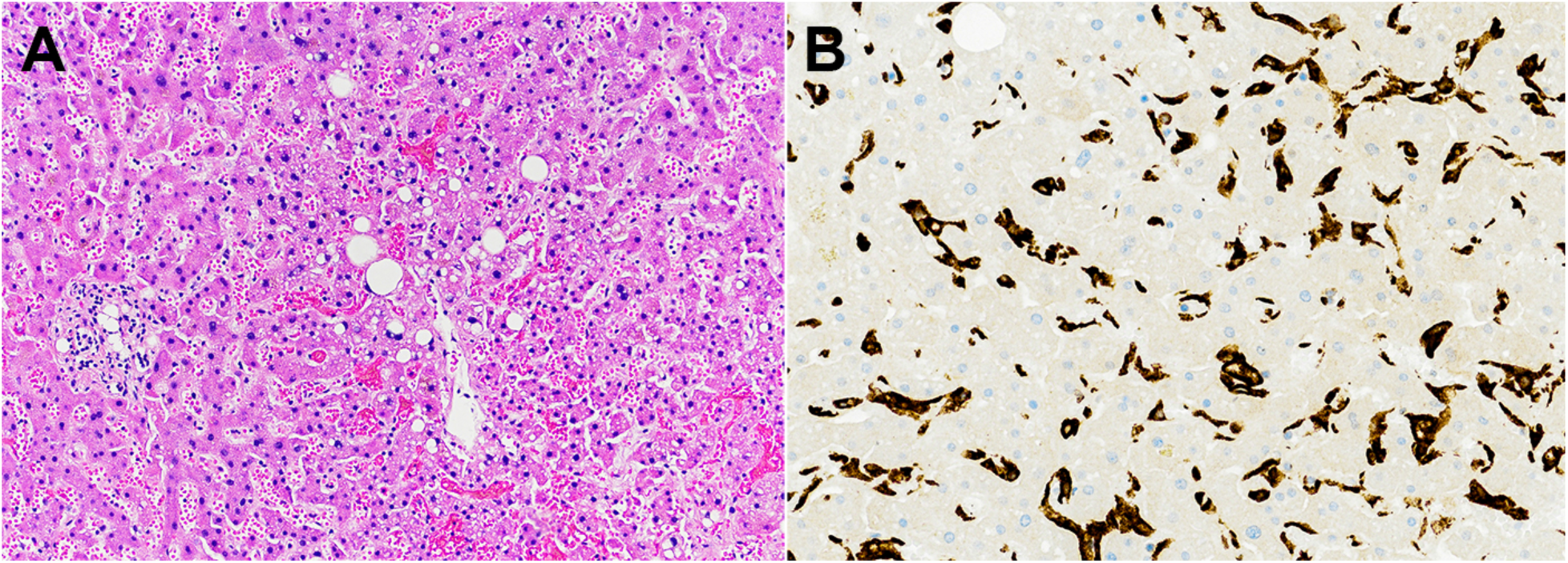
Liver without hemophagocytosis. (A) H&E stain of liver showing mild centrilobular congestion with mild steatosis. (B) CD163 immunohistochemical stain showing mild Kupffer cell hyperplasia without hemophagocytosis.

Bone marrow particles were examined on patient 3 and 4 and these showed trilineage hematopoiesis with left-shifted myeloid hyperplasia. Immunohistochemistry for CD163 showed a mild histiocytic hyperplasia but no hemophagocytosis (images not shown).

### Clinicopathologic Correlation

The presence of hemophagocytosis detected at autopsy was used in conjunction with laboratory findings from pre-mortem laboratory specimens to calculate H-scores (Table 1). Patient 1 had an H-score of 217, meeting the clinicopathologic definition of HLH with extensive hemophagocytosis despite treatment with anakinra. Patient 2 had a partial H-score of 145 as triglyceride level was not available. Given the hemophagocytosis in lymph nodes and enlarged spleen, and progressive rise in ferritin level, we believe this patient likely had HLH. Patient 3 had focal subcapsular hemophagocytosis in a mediastinal lymph node but did not exhibit clinical criteria of HLH with an H-score of 131. Patient 4, who did not show hemophagocytosis in any reticuloendothelial organ, had the lowest H-score of 96. Of note, no patient in our series exhibited hypofibrinogenemia or bicytopenia/pancytopenia. Figure 4 shows the rate of ferritin rise in each of the four patients. Patient 1 and 2 with definite and probable HLH exhibited progressive rise in ferritin levels beginning early in the disease course. Ferritin levels were low throughout most of the disease course of patient 3 and 4 although both received sarilumab early in admission. There was a late rise in ferritin in patient 4 likely reflecting tissue injury resulting from multiple thromboemboli causing pulmonary embolism and stroke the day preceding death. All patients showed elevation of CRP.

**Figure 4.**
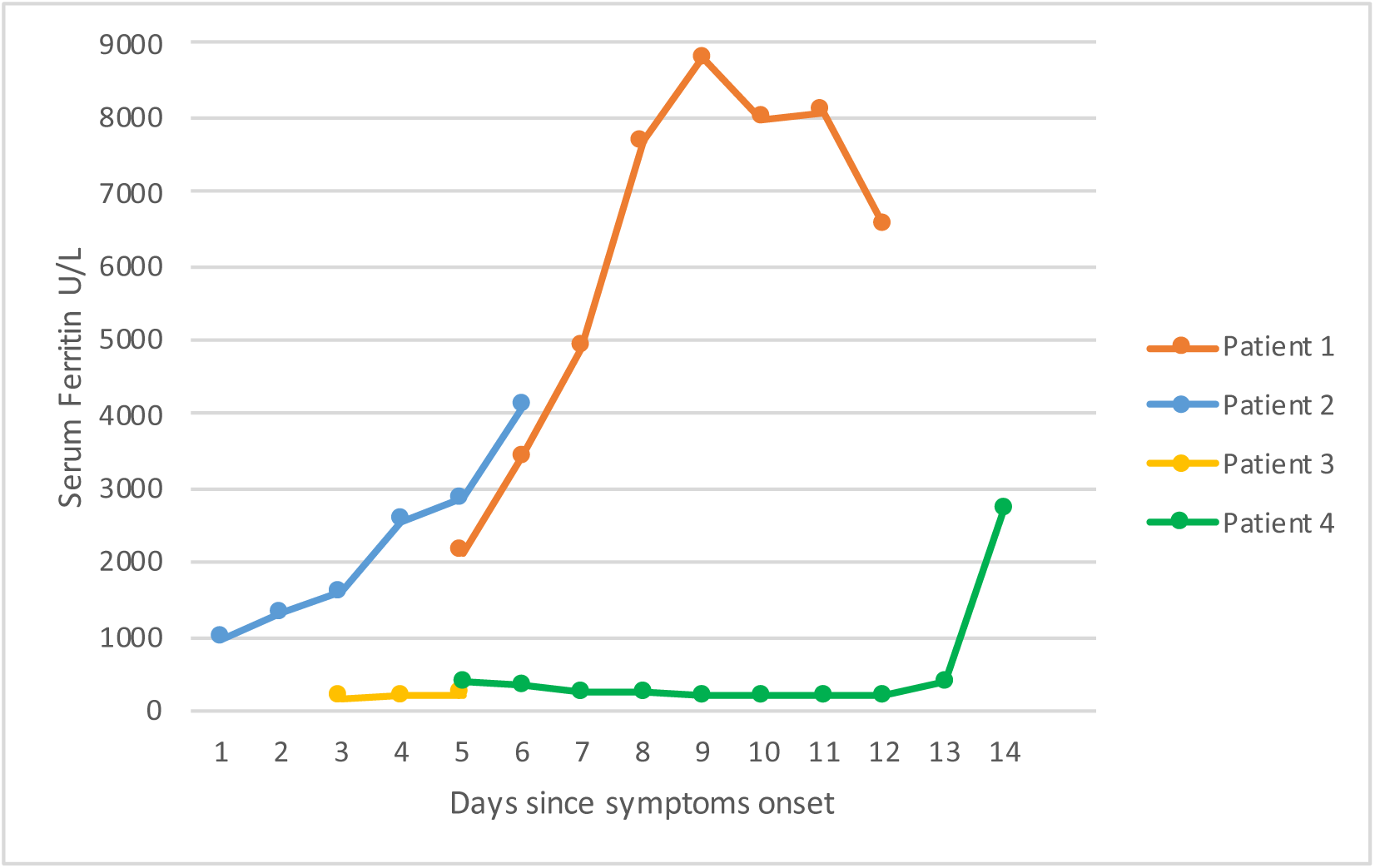
Serum ferritin over disease course.

## Discussion

To date in the pandemic, only a limited number of scientific publications on the histopathologic findings in patients with COVID-19 are available. Most have focused on pulmonary findings either from resections for lung cancer or post-mortem tissue biopsy of lung, heart, and liver.^9–12^ Given the reports of a subset of patients exhibiting clinical features of cytokine storm^5^, we studied our first four autopsies of COVID-19 patients for histologic evidence of hemophagocytosis within the reticuloendothelial organs (lymph node, spleen, bone marrow, and liver) and correlate with clinical and laboratory parameters to detect the syndrome of HLH. We observed one patient with definite HLH and one with probable HLH of the four patients studied.

HLH in adults is a rare life-threatening disease associated with infection, neoplasms, and autoimmune disease.^15^ In the latter situation the term macrophage activation syndrome (MAS) is frequently applied as a subtype of HLH.^16^ Defective granule-mediated cytotoxicity is implicated as the pathophysiologic mechanism resulting in dysregulated antigen presentation and leading to excessive secretion of proinflammatory cytokines referred to as “cytokine storm”. In children this is often related to autosomal recessive mutations in the NK/T-cell cytotoxic pathway termed familial fHLH.^17^ Secondary HLH (sHLH), also called reactive HLH, can present at any age but frequently occurs in patients with underlying diseases affecting immunity including HIV and cancer.^18^

The diagnosis of HLH is based on a constellation of clinical, laboratory and morphologic criteria.^19^ The H-score is a clinical tool which estimates the probability of HLH based upon severity of fever; hepatosplenomegaly; number of cytopenias; elevations in serum ferritin, triglyceride, aspartate aminotransferase (AST); hypofibrinogenemia; and morphologic presence of hemophagocytosis.^13,14^ In a larger cohort of 191 inpatients with COVID-19 in Wuhan, China; non-survivors compared to survivors more frequently had anemia (26% vs. 11%), thrombocytopenia (20% vs. 1%), elevated ALT (48% vs. 24%) and ferritin (96% vs. 71%).^20^ In particular, ferritin levels of 2000 ng/mL or greater were observed in 25% of fatal cases. Alternatively, leukopenia showed the opposite trend being observed in 9% of non-survivors vs. 20% of survivors. The frequency of organomegaly, hypofibrinogenemia, or hypertriglyceridemia is currently not known for COVID-19. In our series, high fever, hyperferritinemia, and hypertriglyceridemia were the most helpful clinical and laboratory finding distinguishing HLH from non-HLH COVID-19 patients with ARDS. Elevated C-reactive protein levels were noted in all four cases, consistent with the cytokine storm that is prevalent in severe COVID-19 infections.^4^ We observed no patient with bicytopenia, pancytopenia, or hypofibrinogenemia as is often seen in other infection-associated HLH but larger studies are required to further characterize whether this is a unique feature of SARS-CoV-2 associated HLH.

Pathologic detection of hemophagocytosis is one criterion used in the diagnosis of HLH. Bone marrow biopsies are the typical antemortem specimen; however, lymph nodes also exhibit characteristic pathologic features.^21^ Some have questioned the specificity of hemophagocytosis as it has also been observed in 64.5% of bone marrow aspirates in patients with sepsis^22^ and rarely within normal bone marrow biopsies.^23,24^ This may be in part explained by the variable definitions of the hemophagocyte; specifically regarding the engulfment of anucleate cells such as erythrocytes and platelets compared to phagocytosis of nucleated cells such as neutrophils, red cell precursors, and lymphocytes. Detailed pathologic studies of bone marrow biopsies in this regard have shown that nucleated cell phagocytosis as well as multiply phagocytosed nucleated cells within single macrophages have higher specificity for the diagnosis of HLH than erythrophagocytosis alone.^25^ Our observation of a high density of macrophages engulfing multiple lymphocytes thus warrants greater consideration than erythrophagocytosis which is observed in a variety of disease states.

Infection-associated sHLH is most often caused by DNA viruses of the Herpesviridae family - Epstein-Barr (EBV) virus, cytomegalovirus (CMV), and Kaposi’s sarcoma-associated virus (KSHV/HHV-8)^15^- which were excluded in our cases. RNA viruses have also been implicated as triggering agents of sHLH, particularly in the epidemic or pandemic setting. In this context, HLH has been observed in a subset of fatal infections with SARS-CoV-1,^26–30^ novel avian-origin influenza A (H5N1)^31–33^ and swine-origin influenza A (H1N1)^34,35^. Intriguingly, among a cohort of 16 fatal H1N1 adult patients, 81% exhibited hemophagocytosis at autopsy and 36% were retrospectively found to harbor heterozygous mutations in familial HLH associated genes using whole-exome sequencing.^36^ Similarly, others have reported that 14% of sporadic adult onset HLH harbor hypomorphic mutations in familial HLH-associated genes.^37^ Taken together, these data may explain why a subset of patients in pandemic settings exhibit a hyper-inflammatory disease course characterized by HLH with cytokine storm. This concept has recently been reviewed and presented as a threshold model for MAS/sHLH.^38^ Prospective molecular studies interrogating these immune abnormalities in individuals with COVID-19 may be useful in predicting which individuals are at greatest risk of cytokine storm.

Herein we report the first documented cases of HLH associated with SARS-CoV-2 whose clinical courses were dominated by ARDS and cytokine storm. High fever, hyperferritinemia, and hypertriglyceridemia were the most useful clinical parameters to identify HLH among our COVID-19 autopsy cohort. Recognition of cytokine storm as a manifestation of sHLH is critical to ensure timely anti-inflammatory treatment concurrent with anti-viral therapy in patients with COVID-19.^5,6^ We note that an etoposide based regimen, typically used to treat EBV-associated HLH, has been used to successfully treat a patient with H1N1-associated HLH who was on extracorporeal membrane oxygenation.^39^ In addition, a variety of targeted approaches against inflammatory cytokines traditionally used in the setting of autoimmune disease-associated MAS have been proposed.^7,8^ Of note, patient 1 who had definitive SARS-CoV-2 associated HLH in our cohort received anakinra (IL-1R antagonist) with persistence of hemophagocytosis at autopsy. Hemophagocytosis but not HLH was seen in one of two patients who received sarilumab (anti-IL-6R mAb). Clinicians should be aware of the signs and laboratory features of HLH, which may develop in a subset of severe SARS-CoV-2 infection. Identification of the hyper-inflammatory clinical phenotype will inform clinical trials of optimal therapy in this life-threatening condition.

## Data Availability

All data is collected by the authors

## Acknowledgments

We would like to acknowledge the excellent technical assistance of Teresa Lima, Emily Aniskovich, Myrtha Constant and Cheryl Spencer.

